# Media coverage and speculation about the impact of the COVID-19 pandemic on suicide: A content analysis of UK news

**DOI:** 10.1101/2022.06.10.22275998

**Authors:** Lisa Marzano, Monica Hawley, Lorna Fraser, Yasmine Lainez, James Marsh, Keith Hawton

**Affiliations:** Faculty of Science and Technology, Middlesex University, The Burroughs, Hendon, London (UK) NW4 4BT; Samaritans, The Upper Mill, Kingston Road, Ewell, Surrey (UK) KT17 2AF; Centre for Suicide Research, Department of Psychiatry, University of Oxford, Warneford Hospital, Oxford (UK) OX3 7JX; Oxford Health NHS Foundation Trust, Warneford Hospital, Oxford (UK) OX3 7JX

**Keywords:** suicide and self-harm, media and journalism, newspaper reporting, media guidelines, COVID-19

## Abstract

**Objectives:** Since the start of the COVID-19 pandemic, there has been much concern and speculation about rises in suicide rates, despite evidence that suicides did not in fact increase in the first year of the pandemic in most countries with real-time suicide data. This public narrative is potentially harmful, as well as misleading, and is likely to be perpetuated by sensational news coverage.

**Method:** We conducted a systematic analysis of UK news coverage (including opinion pieces) on the impact of COVID-19 on suicidality, to examine the content and quality of such reporting as the pandemic developed, and as different coronavirus restrictions were imposed.

**Results:** We identified 372 stories about COVID-19 and suicidality in online and print news between the first UK lockdown (March 2020) and May 2021 (when restrictions were significantly eased in the UK). Throughout this period, over a third of articles (39.2%) and headlines (41.4%) claimed or predicted a rise in suicide, often attributed to feelings of entrapment and poor mental health (especially amongst young people), and fueled by expert commentary and speculation. Almost a third of reports were rated as being of poor overall quality (116, 31.2%), and at least half included no signposting to help and support. However, reporting improved in phases of less stringent COVID-19 restrictions and over time, with later articles and headlines including fewer negative statements and predictions about rises in suicides, and greater reliance on academic evidence.

**Conclusions:** As the longer-term consequences of the pandemic develop, and other national and global events unfold, it is increasingly important that the media, and the wider community of experts shaping its narratives, strive for a positive and evidence-informed approach to news coverage of suicide.

**Strengths and Limitations of this Study:**

- This is the first systematic analysis of UK news coverage on the impact of COVID-19 on suicidality.
- Findings are based on a well-established, evidence-informed news monitoring database.
- However, this analysis cannot account for the impact of news coverage on different audiences, nor for its reach
- Our focus did not extend to broadcasts or other media formats.

## Introduction

News reporting on major health issues can have profound influences on health-related behaviour of the public. One example was the changes in statin use in the UK associated with the widely published controversy in late 2013/early 2014 over the risk-benefit balance of statins for prevention of adverse cardiac events, with an increased number of patients stopping taking statins following this publicity [1]. Another example was the public concern that followed extensive media attention to a report on a small series of children who developed autism after receiving the measles-mumps-rubella (MMR) vaccine. While subsequent studies found no evidence that MMR caused autism, some parents remained concerned about the safety of the vaccine, leading to refusal or delay in its administration [2]. Reporting on health issues can also have positive benefits, an example being the wide reporting of the illness and subsequent death from cervical cancer of the British reality TV star, Jade Goodey, which was followed by an upsurge in cancer screening attendances [3]. More recently, research has shown that the language and tone of public health information can have a significant impact on COVID-19 vaccine beliefs and intentions [4], as well as potentially increasing anxiety and misplaced health-protective behaviours [5].

The nature of news reporting of suicide is also important in relation to influencing suicide risk in the general population [6]. Evidence for this is mainly focussed on news reports of specific suicides, especially deaths of celebrities [7,8] or those using unusual suicide methods [9]. However, general reporting about suicide, including opinion pieces, may also be important, especially those which include predictions about possible rises in suicides. This is likely to be particularly relevant at times of crisis. The COVID-19 pandemic is clearly such a time, with deaths globally so far totalling over 6 million [10] and concerns having been expressed early in the pandemic about the possibility of it having a significant impact on suicide rates [11].

It is also known that positive news reporting, such as people coping with adversity, can have beneficial effects on suicidal behaviour [12]. Other means of broadcasting the need to get help and specific promotion of helplines can also help prevent suicides [13]. Therefore, the way that suicide is addressed in news outlets can be a very important and modifiable influence on suicide in the general population.

In the first year of the pandemic, suicide rates appear not to have risen (and in some cases declined) in most countries for which evidence is available [14,15], including the UK [16]. However, this could change as the longer-term consequences of the pandemic affect the general population, the economy, and high-risk groups, especially if exacerbated by a public narrative dominated by alarming claims and predictions [17].

We previously conducted a study of reports of specific suicides in the UK during the first four months of the pandemic [18]. This showed that many reports made explicit links between suicide and the COVID-19 pandemic in the headline of articles, largely based on based on statements by family, friends or acquaintances. The impact of the pandemic on suicidal behaviour was most often attributed to feelings of isolation, poor mental health and entrapment due to government-imposed restriction. Although rarely of poor overall quality, reporting was biased towards young people, frontline staff and relatively unusual suicides and, to varying degrees, failed to meet recommended standards, such as signposting readers to sources of support. We have now conducted a study of general UK news coverage about suicide and COVID-19 in the first 14 months of the pandemic, including opinion pieces and broader news reports on the impact of the pandemic on suicide rates and trends. The overall aim was to determine the extent to which such news stories and headlines included sensational statements and predictions, and whether they adhered to recommended standards for reporting of suicides.

## Method

### Suicide and COVID-19 news database

Since March 16th 2020 (a week before the first UK national lockdown came into force (on 23rd March 2020)), the suicide prevention charity Samaritans has been monitoring print and online newspaper reports in national and regional British publications which include general statements or speculation about the impact of COVID-19 on suicidality (including suicidal thoughts and behaviour), using a bespoke database developed by the research team. Articles focusing exclusively on mental health and illness are not included in this database, nor are news reports of individual suicides and attempted suicides (the latter have however been analysed as part of a separate study [18]).

All articles in this database are coded by trained researchers and Samaritans’ staff for content, including identification of the article itself (e.g. date, headline and newspaper) and whether explicit statements or predictions about the impact of the pandemic on suicidality (and mental health more generally) are made in the headline and/or the main body of the report. The coding scheme also captures: 1) the extent to which these claims are supported by academic or other evidence; 2) which group or groups are the main focus of each article and headline; 3) which element or elements of the pandemic (and associated challenges and restrictions) are highlighted in headlines, content and images in news stories as contributing to suicidality; 4) any specific issues or concerns in relation to individual articles (e.g. the use of sensational language); and 5) any positive messages being communicated in such reports (e.g. reaching out to loved ones for help). Finally, based on adherence to international recommendations for responsible reporting of suicide during the COVID-19 pandemic [19], and more broadly [20], each news report is also rated for quality (as ‘positive’, ‘neutral’ or ‘negative’) with regards to its headline and overall tone (for further details see [21]), with examples of sensational reporting noted in relation to both.

To ensure that important aspects of the pandemic and its potential influence on suicidal behaviour have been appropriately and comprehensively recorded, the coding scheme was adapted from the ‘Classification of COVID-19 related factors involved in self-harm’ used in an investigation of hospital attendances for self-harm [22]. For other sections of the scheme, codes (where not binary yes/no categories) were derived inductively for content [23] and refined through an iterative process of development and piloting (by three independent raters (MH, YL and JM)).

### Patient and Public involvement

None.

### Data analysis

We analysed the nature and content of all COVID-19 related articles recorded in the Samaritans’ news monitoring database from the week before the first UK national lockdown came into force (on 23rd March 2020) until 17th May 2021, when social distancing restrictions were significantly eased in the UK (at least until December 2021, following the spread of the Omicron variant).

All data are presented as frequencies or percentages (e.g. of items that deviated or not from recommended standards). Variations in the nature and quality of reporting were analyzed using chisquare tests (e.g. in relation to different media types and formats). To capture changes in reporting as the pandemic developed, and associated government-imposed restrictions were imposed, we identified four main ‘restriction phases’ during the 14-month period of our study, based on the official timeline of UK government coronavirus lockdowns and measures [24]. These included:

- Two phases of lockdown and greater social distancing measures:

◦ Phase 1, from 23rd March to 22nd June 2020
◦ Phase 3, from 31st October 2020 to 22nd February 2021
- Two periods of less stringent restrictions:

◦ Phase 2, from 23rd June to 30th October 2020
◦ Phase 4, from 23rd February to 17th May 2021.

## Results

Between 16th March 2020 and 17th May 2021, we identified 372 articles in the British news focusing or speculating on the impact of the pandemic on suicidal behaviour. Of these, approximately half were online reports (193, 51.9%), and appeared in local and regional news (180, 48.4%; as opposed to tabloids (95, 25.5%) or broadsheets (97, 26.1%)). Nearly one in ten print articles were located in prominent positions within the paper (in 16/177 print articles these were on pages 1 to 3).

Although there were no COVID-19 related reports in the first nine days of monitoring, most weeks thereafter saw the publication of several stories about the pandemic and suicidality (weekly median=5), with a peak in July 2020 (Figure 1).

**Figure 1.**
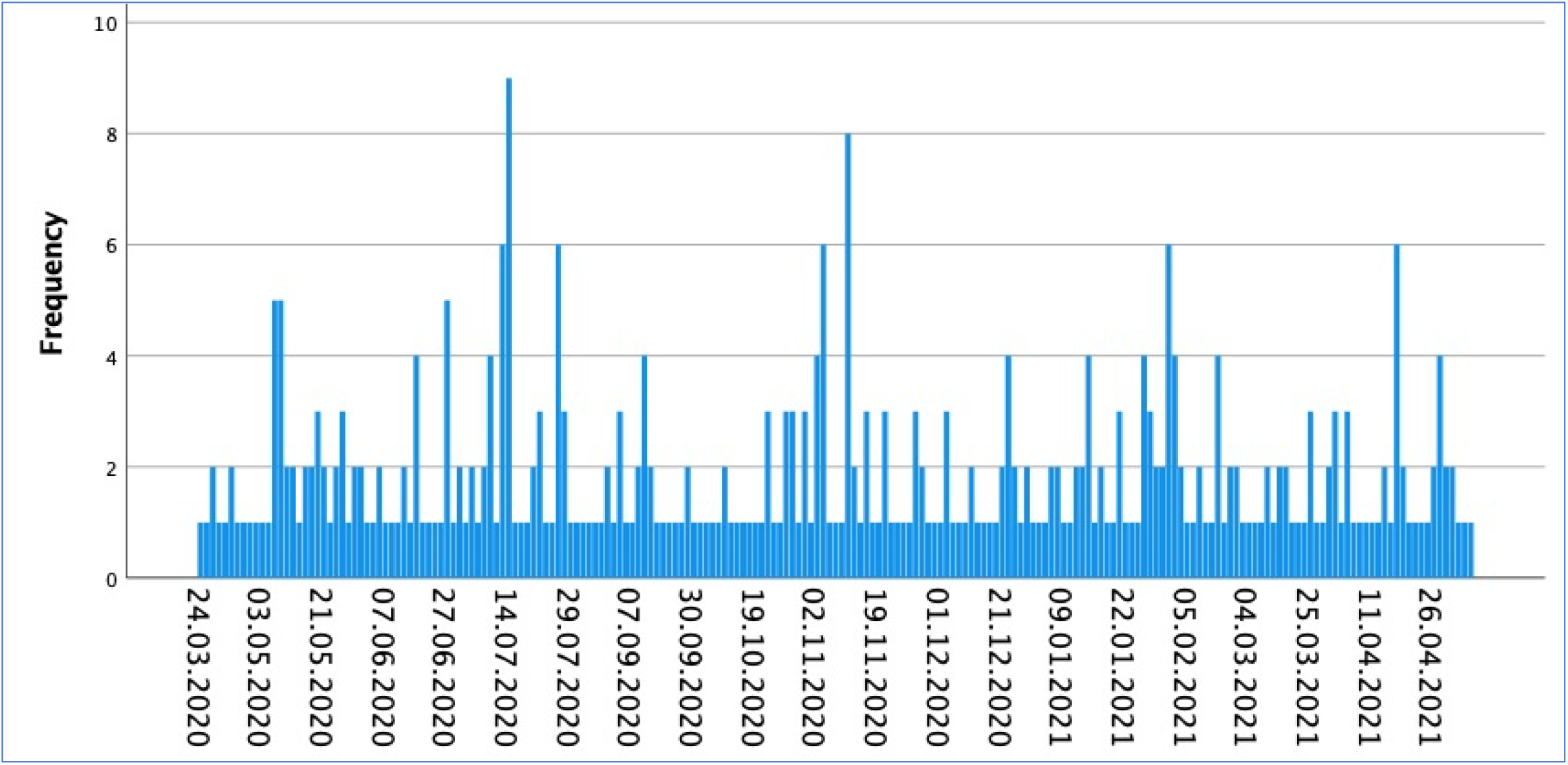
Daily coverage about the impact of the COVID-19 pandemic on suicide in national and regional British news (23rd March 2020 to 17^th^ May 2021)

There were proportionally more reports about the impact of the pandemic on suicidal behaviour at times of greater social distancing restrictions (201, 54% of all articles were published in the 214 days of stricter government-imposed restrictions (phases 1 and 3) vs. 171 (46.0%) in the 214 days of phases 2 and 4), particularly in the October 2020 to February 2021 lockdown period (on average, more than one article about suicide and the pandemic was published daily in phase 3).

### COVID-19 and suicide: focus and content of news headlines and reports

Newspaper reports were selected on the basis of their focusing on, or making reference to the potential or alleged impact of the pandemic on suicide rates. Indeed, throughout the study period, most stories made strong and direct links between the pandemic and suicide (332, 89.2%), and most headlines included an explicit focus on suicide (238, 64.0% vs. 98, 26.3% focusing on mental health more generally) and on COVID-19 (211, 56.7%), especially at the start of the pandemic (51/66, 77.3%) (Tables 1 & 2). Particularly in earlier phases of the pandemic, location/region/country details were commonly included in story titles (in 77/120 such headlines (64.2%) the focus was on specific UK regions (n=35), counties (n=16) or cities (n=7), or the UK as a whole (n=19); vs. 44 headlines focusing on other countries, especially the US (n=14), Japan (n=13) and Australia (n=9)).

**Table 1:** Article headlines about COVID-19 and suicidality in online and print British news, over the first four phases of the pandemic (16th March 2020-17th May 2021)

|  | **16<sup>th</sup> March to**<br>**22<sup>nd</sup> June 2020** |  | **23<sup>rd</sup> June to**<br>**30<sup>th</sup> October 2020** |  | **31<sup>st</sup> October 2020 to**<br>**22<sup>nd</sup> February 2021** |  | **23<sup>rd</sup> February to**<br>**17<sup>th</sup> May 2021** |  |
| --- | --- | --- | --- | --- | --- | --- | --- | --- |
| **HEADLINE** | **n/66** | **(%)** | **n/108** | **(%)** | **n/135** | **(%)** | **n/63** | **(%)** |
| **Explicit focus on suicide** | 39 | 59.1% | 72 | 66.7% | 84 | 62.2% | 43 | 68.3% |
| **Explicit focus/terminology relating to COVID-19** | 51 | 77.3% | 56 | 51.9% | 69 | 51.1% | 35 | 55.6% |
| **Focus on specific location/region/country** | 36 | 55.5% | 30 | 27.8% | 35 | 25.9% | 19 | 30.2% |
| **Focus on suicide prevention/call for action** | 11 | 16.7% | 14 | 13.0% | 5 | 3.7% | 3 | 4.8% |
| **Sensational tone or language** | 28 | 42.4% | 34 | 31.5% | 76 | 56.3% | 28 | 45.2% |
| **Includes negative prediction (any)** | 14 | 21.2% | 28 | 25.9% | 27 | 20.0% | 9 | 14.3% |
| • About suicide | 9 | 13.6% | 13 | 12.0% | 15 | 11.1% | 4 | 6.3% |
| • About population mental health | 4 | 6.1% | 13 | 12.0% | 9 | 7.7% | 5 | 7.9% |
| • About both suicide and mental health | 1 | 1.5% | 1 | 0.9% | 3 | 2.2% | 0 | 0.0% |
| • Other | 0 | 0.0% | 1 | 0.9% | 0 | 0.0% | 0 | 0.0% |
| **States increase in:** |  |  |  |  |  |  |  |  |
| • Suicide rates/numbers | 12 | 18.2% | 20 | 18.5% | 37 | 27.4% | 11 | 17.7% |
| • Self-harm/suicidal behaviour | 1 | 1.5% | 3 | 2.8% | 0 | 0.0% | 0 | 0.0% |
| • Suicidal thoughts/expressions of suicidality | 2 | 3.0% | 5 | 4.6% | 13 | 9.6% | 1 | 1.6% |
| • Help-seeking for suicidality | 4 | 6.1% | 1 | 0.9% | 7 | 5.2% | 2 | 3.2% |
| • Mental health problems | 3 | 4.5% | 9 | 8.3% | 24 | 17.8% | 13 | 21% |
| • Other (e.g. drugs) | 1 | 1.5% | 0 | 0.0% | 2 | 1.5% | 3 | 4.8% |
| **States decrease or no change in:** |  |  |  |  |  |  |  |  |
| • Suicide rates/numbers | 3 | 4.5% | 6 | 5.6% | 7 | 5.2% | 18 | 29.0% |
| • Self-harm/suicidal behaviour<br>• Mental health problems | 0<br>0 | 0.0%<br>0.0% | 0<br>1 | 0.0%<br>0.9% | 0<br>0 | 0.0%<br>0.0% | 1<br>1 | 1.6%<br>1.6% |
| **Single event as headline ‘hook’** | 2 | 3.0% | 10 | 9.3% | 19 | 14.1% | 3 | 4.8% |
| **Includes statement/quote by:** |  |  |  |  |  |  |  |  |
| • Academic<br>• Government/politician<br>• Charity/Third sector<br>• Clinician/NHS<br>• Emergency services<br>• Person bereaved by suicide<br>• Person with lived experience of suicidality<br>• Unspecified expert/s<br>• Other | 3<br>0<br>6<br>10<br>0<br>0<br>0<br>3<br>2 | 4.5%<br>0.0%<br>9.1%<br>15.2%<br>0.0%<br>0.0%<br>0.0%<br>4.5%<br>3.0% | 4<br>7<br>6<br>9<br>1<br>1<br>2<br>0<br>3 | 3.7%<br>6.5%<br>5.6%<br>8.3%<br>0.9%<br>0.9%<br>1.9%<br>0.0%<br>2.8% | 2<br>3<br>2<br>6<br>0<br>3<br>2<br>0<br>5 | 1.5%<br>2.2%<br>1.5%<br>4.4%<br>0.0%<br>2.2%<br>1.5%<br>0.0%<br>3.7% | 0<br>0<br>2<br>2<br>0<br>0<br>1<br>0<br>0 | 0.0%<br>0.0%<br>3.2%<br>3.2%<br>0.0%<br>0.0%<br>1.6%<br>0.0%<br>0.0% |

**Table 2:**
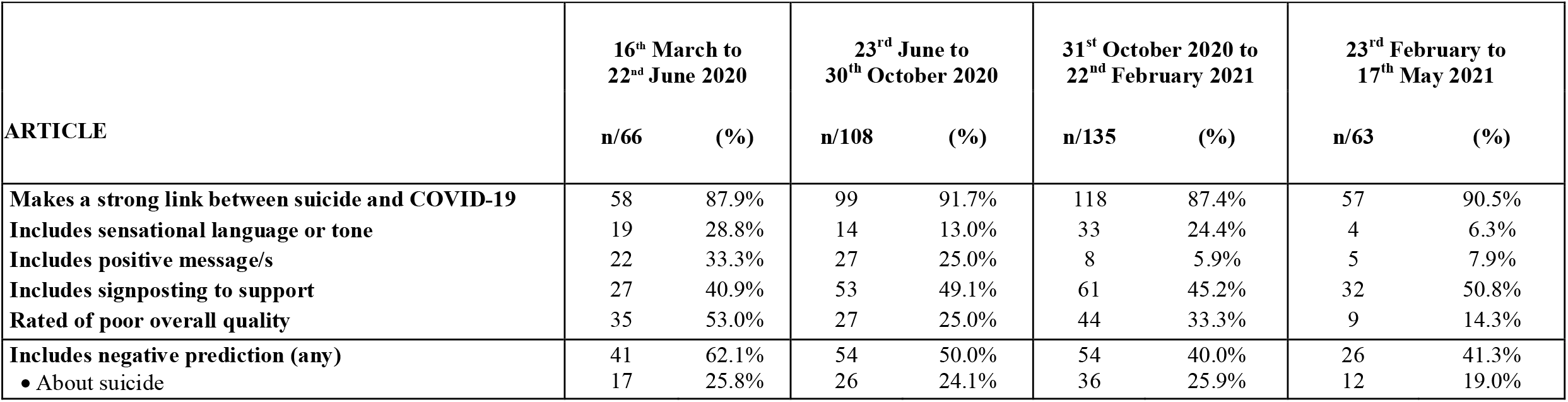
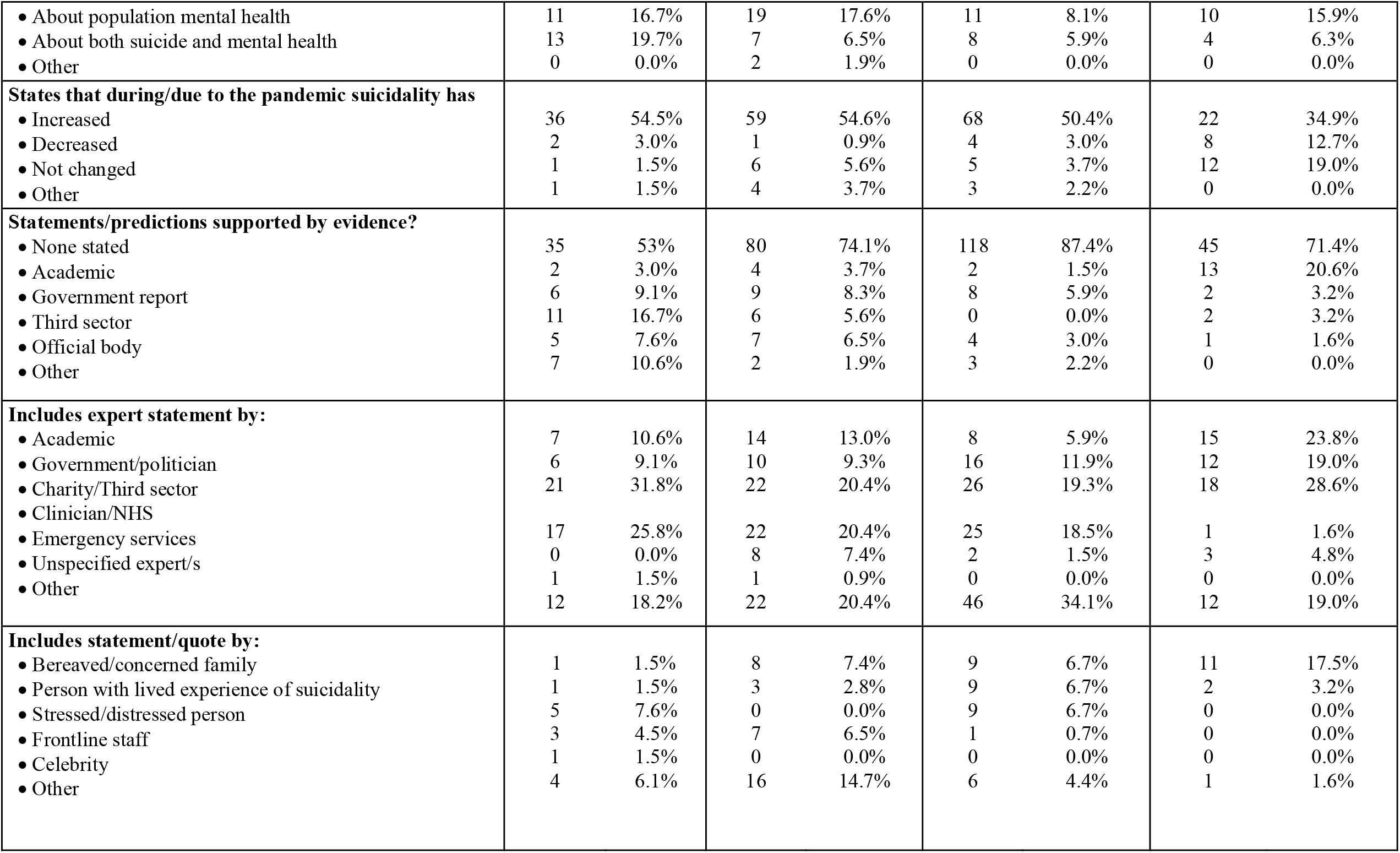
Content and quality of articles about COVID-19 and suicidality in online and print British news, over the first four phases of the pandemic (16th March 2020-17th May 2021)

In some cases, headlines referred to individual events or alleged clusters as a ‘hook’ into broader news reports on the impact of the pandemic on suicide rates and trends more generally. More often, headlines featured statements by experts (including clinicians, mental health charities, academics and politicians) about the impact of the pandemic on mental health and suicide rates (Table 1). Expert speculation was especially dominant in news headlines at the start of the pandemic, featuring in over a third of headlines between March and October 2020 (24/66, 36.3% in phase 1 and 33/108, 30.6% in phase 2 vs. 23/135, 17.0% in phase 3 and 5/63 8.0% in phase 4), thus becoming an important influence on media narratives at a time when official evidence about the impact of COVID-19 on suicide rates was lacking.

### COVID-19 and suicide: focus and content of news headlines and reports

Newspaper reports were selected on the basis of their focusing on, or making reference to the potential or alleged impact of the pandemic on suicide rates. Indeed, throughout the study period, most stories made strong and direct links between the pandemic and suicide (332, 89.2%), and most headlines included an explicit focus on suicide (238, 64.0% vs. 98, 26.3% focusing on mental health more generally) and on COVID-19 (211, 56.7%), especially at the start of the pandemic (51/66, 77.3%). Particularly in earlier phases of the pandemic, location/region/country details were commonly included in story titles (in 77/120 such headlines (64.2%) the focus was on specific UK regions (n=35), counties (n=16) or cities (n=7), or the UK as a whole (n=19); vs. 44 headlines focusing on other countries, especially the US (n=14), Japan (n=13) and Australia (n=9)).

In some cases, headlines referred to individual events or alleged clusters as a ‘hook’ into broader news reports on the impact of the pandemic on suicide rates and trends more generally. More often, headlines featured statements by experts (including clinicians, mental health charities, academics and politicians) about the impact of the pandemic on mental health and suicide rates (Table 1). Expert speculation was especially dominant in news headlines at the start of the pandemic, featuring in over a third of headlines between March and October 2020 (24/66, 36.3% in phase 1 and 33/108, 30.6% in phase 2 vs. 23/135, 17.0% in phase 3 and 5/63 8.0% in phase 4), thus becoming an important influence on media narratives at a time when official evidence about the impact of COVID-19 on suicide rates was lacking.

At the start of the pandemic, almost one in four headlines focused on the effect of COVID-19 on suicide rates in the general population (Figure 2). In subsequent restriction phases, children and young people were more often the focus of news headlines, featuring in approximately 1 in 6 titles. In the articles themselves, children and young people were also a common focal point (on average, just under 20% of news reported specifically, and sometimes exclusively, on the impact of the pandemic on younger age groups), though most stories centred around general population effects (65.3%). Other groups (e.g. older people, keyworkers, and people with mental health issues) were rarely the main focus of the news stories and headlines.

**Figure 2a.**
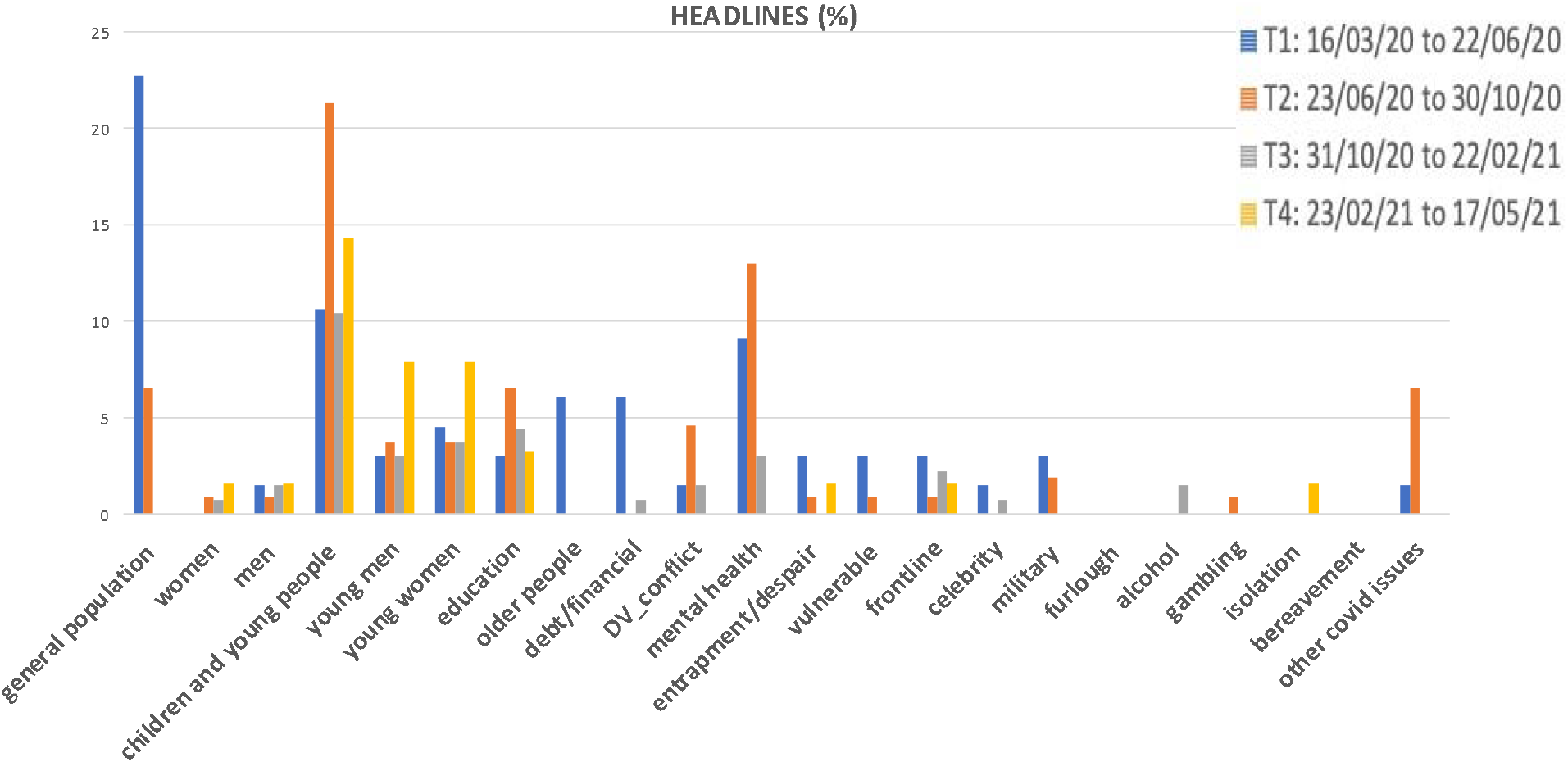
News coverage of COVID-19 and suicidality: Proportion of headlines focusing on specific issues and populations.

**Figure 2b.**
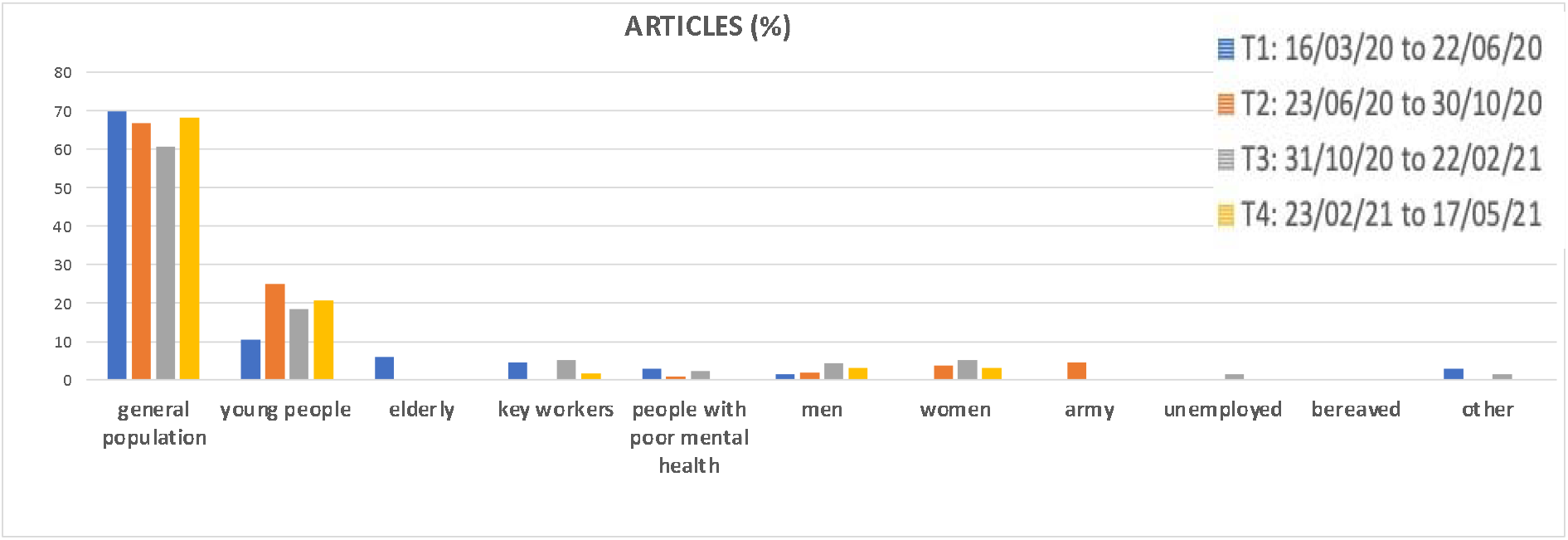
News coverage of COVID-19 and suicidality: Proportion of articles focusing specific issues and populations.

In news reports, as well as in headlines, the link between suicide and COVID-19 was often discussed in fairly generic terms (rather than ascribed to specific issues such as infection fears, bereavement or disruption to schooling) (Figure 3). However, especially after the initial lockdown period, feelings of entrapment and poor mental health (both worsening mental health and emerging, pandemic-related mental health issues) became common themes in news stories and headlines about COVID-19 and suicide. Financial and employment issues, and feelings of isolation due to government restrictions, were cited less often, and very few headlines or reports discussed the impact of COVID-19 on suicidality with reference to increases in domestic violence, alcohol and drugs use/abuse and housing issues, or the stresses and trauma of working on the front-line during the pandemic.

**Figure 3a.**
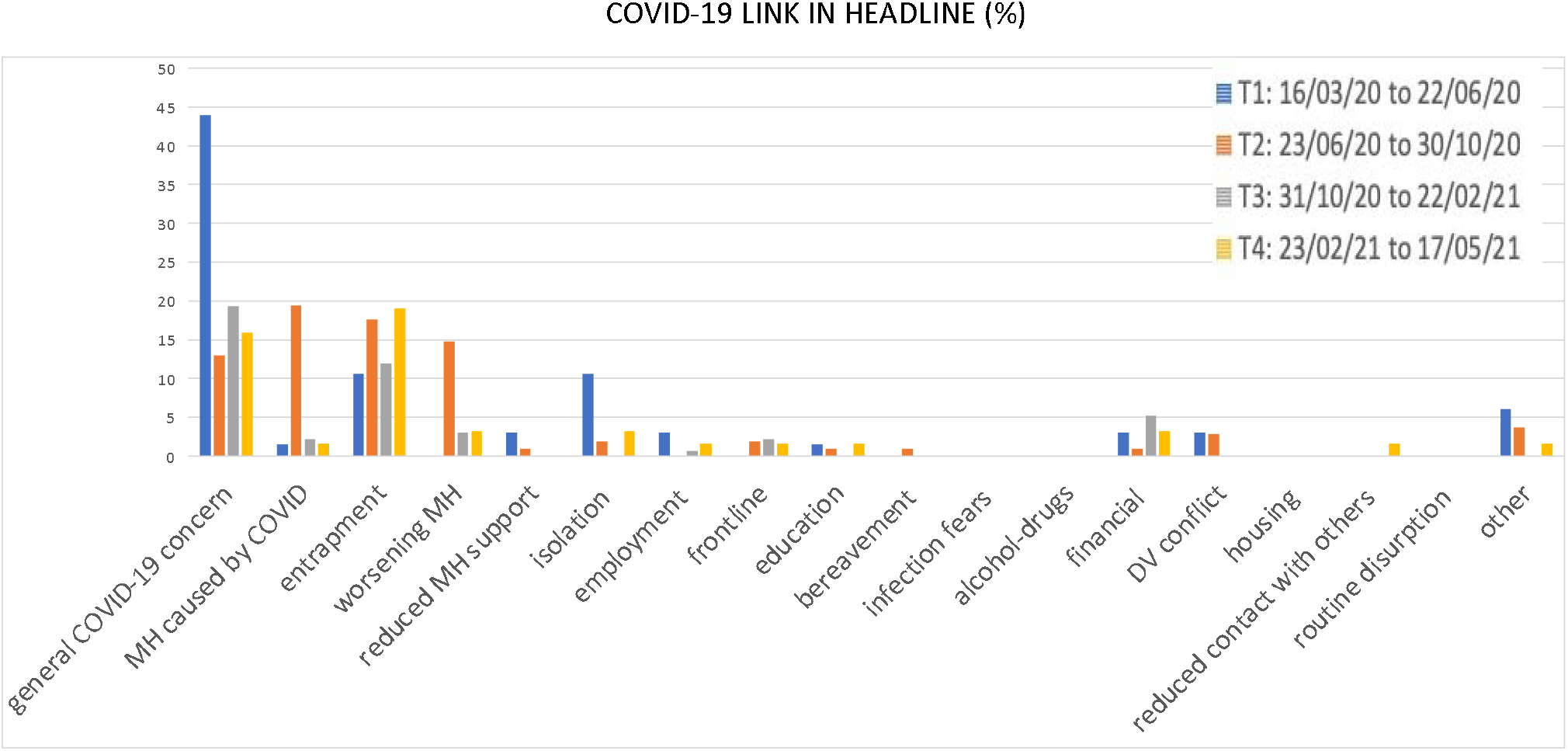
Suggested COVID-19 related influences on suicidal behaviour in news headlines.

**Figure 3b.**
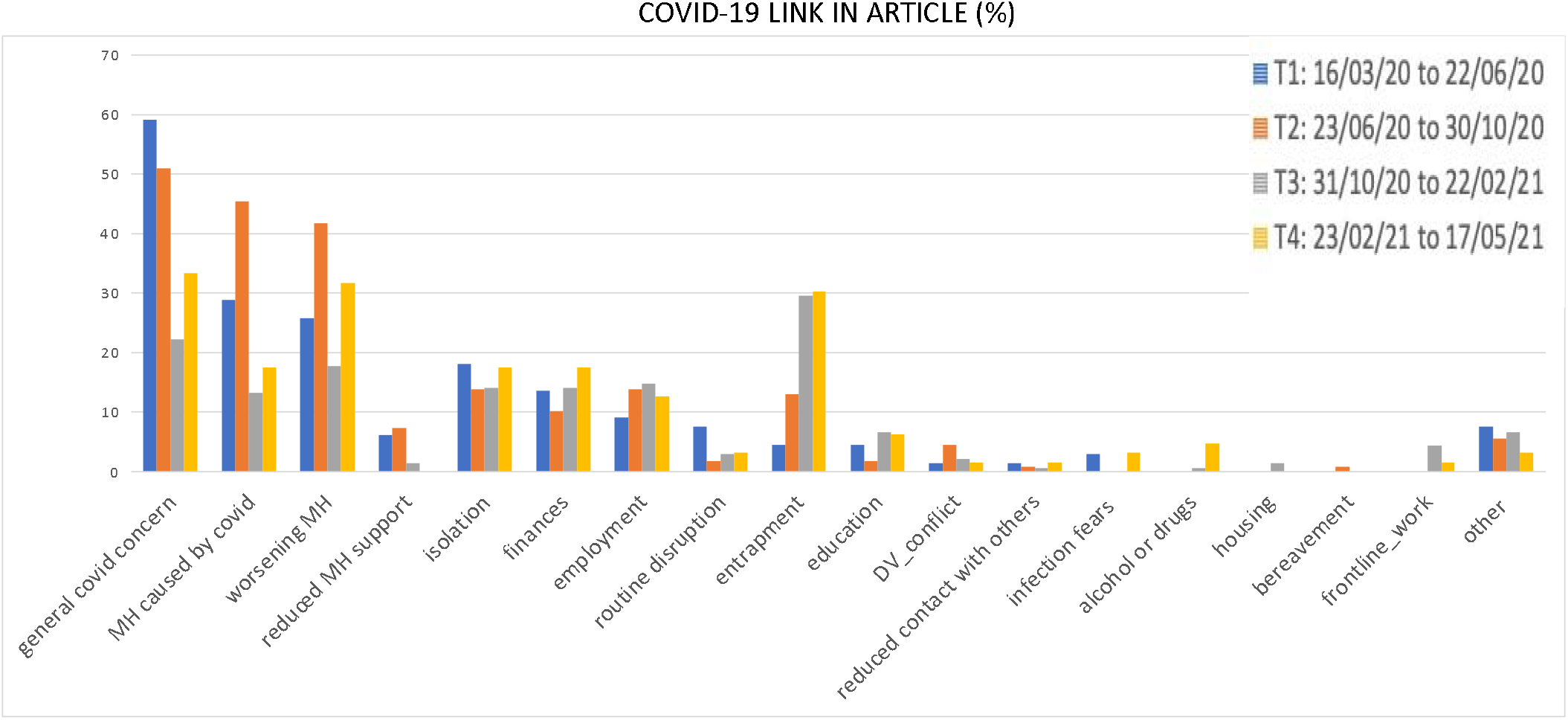
Suggested COVID-19 related influences on suicidal behaviour in news ARTICLES.

Of note was the low number of reports discussing specific suicide methods, including in headlines (n=5).

### COVID-19 imagery in news reports about suicide and the pandemic

Relatively few reports included images relating to the pandemic (66, 17.7%) (e.g., medical settings, equipment or staff (22, 5.9%) or COVID-19 infection graphs (12, 3.2%)), with some including images of individuals who had attempted or died by suicide (19, 5.1%) or persons connected to those who had died (6, 1.6%). Very few included images of suicide locations (2, 0.6%) and none of specific suicide methods. Covid-related imagery was more common in phases of greater social distancing restrictions (46/201, 22.9% vs. 20/171, 11.7% of reports during phases 2 and 4, X^2^(1)=7.927, p<0.05).

### What, if any, is the impact of the pandemic on suicidality?

A third of headlines asserted an increase in suicidality due to COVID-19 and associated restrictions (119, 32%), of which 80, 67.2% were about rises in suicide rates (vs. 4, 3.4% non-fatal suicidal behaviour, 21, 17.6% suicidal ideation or 14, 11.8% help-seeking for suicidality). An additional 12.4% predicted a rise in suicides (Table 1). At times of greater restrictions, more headlines stated or predicted an increase in suicidality (92/201, 45.8% vs. 54/171, 31.6% when restrictions were less stringent; X^2^(1)=7.805, p=0.005).

Of note is the larger proportion of headlines reporting a decrease in suicidality in the final study period (18/60, 30.0%) compared to earlier phases (ranging from 4.5% to 5.6%, X^2^(3)=36.903, p<0.001), particularly in online news (25/195, 12.8% vs. 9/177, 5.1% print articles; X^2^(1)=5.438, p<0.05). This also reflects the growing proportion of articles stating that rates of suicide and suicidality had decreased or remained stable since the start of the pandemic (20/63, 31.7% in phase 4 vs. 4.6% in phase 1, 6.5% in phase 2 and 6.7% in phase 3; X^2^(3)=36.586, p<0.001).

Nonetheless, such articles continued to be less common than reports stating or predicting a rise in suicides, in both national and regional publications. Overall, around 50% of articles appearing in print and online broadsheet news (48/97), tabloids (48/95) and regional press (89/180) reported an increase in suicidality during the pandemic (Table 2; this proportion was slightly lower between February and May 2021 (34.9%) than in earlier phases of the pandemic, but not sufficiently to reach statistical significance (X^2^(3)=7.415, p=0.60)). A smaller but considerable proportion of stories included negative predictions or warnings about such rises (122/372, 32.8% over the study period). The proportion of articles asserting or predicting rises in suicidality was slightly higher at times of greater restrictions, but this difference was not statistically significant (91/201, 45.3% vs. 63/171, 36.8% when restrictions were less stringent, X^2^(1)=2.708, p=0.1).

### Reliance on official evidence

The proportion of news reports citing official evidence to support the statements and speculations made was generally low, and decreased over time (Table 2). Throughout the study period, most stories making a direct link between the pandemic and suicide rates included no evidence to support this (252/332, 75.9%), including when stating or predicting increases in suicide as a result of the pandemic (106/154, 68.8%). This was especially the case in print news compared to online news reports (152/177, 85.9% vs. 126/195, 64.6%; X^2^(1)=22.208, p<0.0001). Reliance on academic sources (and statements by academic experts) increased over time, but was overall limited (only 21 reports cited academic evidence; in 9 cases to support an increase in suicidality and in 11 to state that there had been no change in suicidality). Eleven articles included links to research evidence (3 separate studies)). Other evidence sources (including government, third sector and official bodies reports/statements) were also seldom cited, but were more often used to support an increase in suicidality as a result of the pandemic. Only three articles explicitly questioned the available evidence, or discussed the paucity of evidence on the impact of the pandemic on suicidal behaviour.

### Quality of reporting

Almost a third of reports were rated as being of poor overall quality (116, 31.2%). Reporting was generally poorer in periods of greater social distancing restrictions (80/201, 39.8% vs. 36/171, 21.2%, X^2^(2)=15.159, p<0.001), particularly at the start of the pandemic, and in tabloid press (42.95, 44.2% of tabloid stories were rated negatively vs. 26/97, 26.8% of broadsheet and 48/180, 26.7% of regional reports, X^2^(2)=12.216, p<0.05). Over a quarter of tabloid news stories included examples of sensational language (27, 28.4% vs. 16, 16.5% of broadsheet and 27, 15.0% of regional news; X^2^(2)=7.795, p<0.005), for instance references to a “spiralling”, “skyrocketing”, “dizzying rise in suicides” or a “tsunami”/“tidal wave” of “a myriad of mental health problems”.

Examples of sensational language and tone were even more common in headlines (both in print and online news), especially in phases of greater social distancing restrictions (104/201, 51.7% vs. 62/170, 36.5% when restrictions were relatively less stringent; X^2^(1)= 8.687, p<0.05).

### Signposting and positive messages

Only half or fewer reports included signposting to help and support (Table 2). Print articles were especially likely to omit such detail (116/177, 65.5% vs. 83/195, 42.6% online news; X^2^(1)=19.682, p<0.001). Although of comparable overall quality, print reports were also less likely to include messages deemed to be positive (10/177, 5.6% vs. 46/195, 23.6%; X^2^(1)=23.352, p<0.0001), for instance about suicides (and suicide increases) not being inevitable (19, 5.1% vs. 0 in print news). Nonetheless, there were examples in both print and online news of stories emphasizing the importance of help-seeking (26, 7%) and reaching out to loved ones (4, 1.1%); and of talking about suicide (3, 0.8%) and mental health more generally (10, 2.7%). Such messages appeared in a quarter of news stories reporting or predicting increases in suicidality (41/154, 26.6% vs. 21/218, 9.6% of stories not claiming or predicting such rise; X^2^(1)=18.757, p<0.0001), especially at the start of the pandemic, when news headlines were also more likely to include a focus on suicide prevention initiatives, calls for action and help-seeking (Table 1).

## Discussion

We analysed news coverage of the impact of the pandemic on suicidality in the UK between March 2020 (when the first UK lockdown begun) to May 2021 (when restrictions were significantly eased under the UK Government’s ‘roadmap for the lifting the lockdown’ [24]), to examine the content and quality of such reporting as the pandemic developed, and as different pandemic-related lockdowns and measures were imposed. Our findings suggest that stories about the pandemic and suicide were more frequent and of poorer quality in phases of greater social distancing restrictions. Given the potential impact of such reporting on population level suicide risk, it is especially important that news coverage of suicide is as responsible and balanced as possible when individuals at risk of suicide and the wider public are likely to be experiencing increased distress and mental health difficulties [25–27]. Evidence-based guidelines and advice are available to the media, members of the research community, health and third sector experts on how to safely cover the topic of suicide (via *<u>mediaadvice@samaritans</u>*.*<u>org</u>* and [20]).

Of particular concern was the high proportion of articles (39.2%), including headlines (41.4%), throughout the pandemic which asserted and/or predicted rises in suicide. This was especially true in tabloid news and in headlines, in which this was often portrayed as a “dizzying rise” (across all media types and formats, 44.7% of titles and 18.8% of articles included examples of sensational language), despite no official evidence to support such claims [15,16]. Indeed, although references to evidence (particularly from academic research) increased as the pandemic developed, these remained sparce throughout. This is perhaps unsurprising given the objective paucity of timely evidence on the impact of the pandemic on suicidal behaviour. The absence of an established ‘real-time surveillance system’ to monitor national suicide trends (without the considerable delays which accompany the publication of official suicide statistics [28]) leaves a vacuum of reliable information for the media (and indeed others) to draw upon. In this context, expert speculation about rises in suicide and alarmist comments about declining population mental health can fuel unhelpful media narratives about suicide increases.

A balanced approach to expert commentary and evidence-informed news coverage are important to avoid fuelling harmful myths and misinformation [29]. These could lead to imitative behaviours, and also misguide public opinion about suicide risk. In the news reports, feelings of entrapment and poor mental health were portrayed as key in relation to suicide (see also [18]), especially after the initial lockdown phase. This is consistent with emerging evidence on the factors contributing to hospital presentations following self-harm during lockdown restrictions in England [22]. However, surprisingly few articles or headlines highlighted how specific groups may be affected by the pandemic (e.g. bereaved individuals, frontline staff, women affected by domestic violence, or individuals suffering financial, addiction, housing or physical health issues, including ‘long COVID’).

Also, whilst most news stories focused on the wider public, there was a disproportionate focus on children and young people relative to their actual involvement in suicide [30], as previously observed in news coverage of individual suicides (before [21] and during the pandemic [18]). Given that suicide rates in this group have been increasing for over ten years (whilst remaining lower than for any other age group in England ([31,32]), this is perhaps understandable - but nonetheless concerning. Young people have been shown to be especially susceptible to the influence of news reports on suicidal behaviour [33].

On a positive note, we identified a gradual improvement in reporting over time. Coinciding with the publication of academic papers showing that suicides had not in fact risen at the start of the pandemic [15,16], news articles in the latest restriction period examined (February to May 2021) included fewer claims and predictions about rises in suicides, more headlines stating that suicide rates had decreased or remained stable during the pandemic, and greater reliance on academic evidence. This was particularly the case in online news which, possibly because less constrained in relation to space/word count than printed newspapers, also included more messages deemed to be positive (e.g. about suicide being preventable), and more signposting to sources of support.

Of note, however, was the inclusion of other messages deemed to be positive (e.g. calls for increased help and help-seeking for suicidality) in news reports predicting or stating increases in suicidality. Clearly, the relationship between story newsworthiness and quality of reporting is a complex one. The impact of such reports on different audiences is also likely to be complicated, and requires separate investigation.

### Strengths and Limitations

This study is the first systematic analysis of UK news coverage on the impact of COVID-19 on suicidality at a time of heightened concern and speculation over the effects of the pandemic and associated restrictions on mental health, in academic literature [34,35] and public narrative [17]. Our findings are based on a well-established, evidence-informed media monitoring database which captures media reports of suicides and attempted suicides in the UK [36]. This was carefully adapted for use with generic reports (a separate study was carried out to examine the quality and content of UK news coverage of individual suicides and attempted suicides in the first 4 months of the COVID-19 pandemic [18]). However, our database may not capture all relevant reports, or all important aspects of reporting. Also our focus did not extend to broadcasts or other media formats, and we did not estimate the impact or reach of different news reports (including the circulation rate of different story types, in news as well as social media channels).

### Conclusions

Sensational predictions and unsubstantiated claims of suicide rises were common features of news reports about the impact of the pandemic on suicidality in the UK over the first 14 months of the pandemic, particularly at times of greater social distancing restrictions. However, reporting improved over time, with a growing number of news stories and headlines reflecting observational data that suicide rates did not in fact increase at the start of the pandemic in England [16] or in most other countries for which such evidence is available [14,15].

Further research is needed to assess the quality, content and impact of general reporting about suicide, including opinion pieces, as the longer-term consequences of the pandemic develop [37], and other national and global events unfold (not least the UK energy crisis and the war in Ukraine). A more nuanced understanding of the impact and reach of news stories claiming or predicting rises in suicides is needed, including in relation to their potential benefits [6,38] and unintended consequences for different audiences, and in different types of media (e.g. on social media, and in public awareness campaigns and broadcasts given by members of the research community, health and third sector experts [29]).

## Data Availability

All data produced in the present study are available upon reasonable request to the authors

## Ethics Statement

Not applicable/No human participants included.

## Contributorship statement

All persons who meet authorship criteria are listed as authors, and all authors certify that they have participated sufficiently in the work to take public responsibility for the content, including participation in the concept, design, analysis, writing, or revision of the manuscript.

In particular: KH, LM, MH & LF conceived the study and developed the basis for the protocol.

All authors reviewed and refined the study protocol and coding scheme. MH, JM and YL coded the newspaper articles.

LM analysed the data and, with KH, wrote the initial version of the manuscript.

All authors contributed to the interpretation of results and revision of the article.

All authors approved the final article.

## Data sharing statement

The data that support the findings of this study are available on request from the corresponding author, LM

## Acknowledgments

The authors are very grateful to Jacqui Morrissey, Assistant Director, Research and Influencing at Samaritans, for her invaluable support and feedback.

